# Enhanced Patient Portal Engagement Associated with Improved Weight Loss Outcomes in Post-Bariatric Surgery Patients

**DOI:** 10.1101/2024.01.20.24301550

**Authors:** Xinmeng Zhang, Kaidi Kang, Chao Yan, Yubo Feng, Simon Vandekar, Danxia Yu, S. Trent Rosenbloom, Jason Samuels, Gitanjali Srivastava, Brandon Williams, Vance L. Albaugh, Wayne J. English, Charles R. Flynn, You Chen

## Abstract

**Background:** Bariatric surgery is an effective intervention for obesity, but it requires comprehensive postoperative self-management to achieve optimal outcomes. While patient portals are generally seen as beneficial in engaging patients in health management, the link between their use and post-bariatric surgery weight loss remains unclear.

**Objective:** This study investigated the association between patient portal engagement and postoperative body mass index (BMI) reduction among bariatric surgery patients.

**Methods:** This retrospective longitudinal study included patients who underwent Roux-en-Y gastric bypass (RYGB) or sleeve gastrectomy (SG) at Vanderbilt University Medical Center (VUMC) between January 2018 and March 2021. Using generalized estimating equations, we estimated the association between active days of postoperative patient portal use and the reduction of BMI percentage (%BMI) at 3, 6, and 12 months post-surgery. Covariates included duration since surgery, the patient’s age at the time of surgery, gender, race and ethnicity, type of bariatric surgery, severity of comorbid conditions, and socioeconomic disadvantage.

**Results:** The study included 1,415 patients, mostly female (80.9%), with diverse racial and ethnic backgrounds. 805 (56.9%) patients underwent RYGB and 610 (43.1%) underwent SG. By one-year post-surgery, the mean (SD) %BMI reduction was 31.1% (8.3%), and the mean (SD) number of patient portal active days was 61.0 (41.2). A significantly positive association was observed between patient portal engagement and %BMI reduction, with variations revealed over time. Each 10-day increment of active portal use was associated with a 0.57% ([95% CI: 0.42- 0.72], *P* < .001) and 0.35% ([95% CI: 0.22- 0.49], *P* < .001) %BMI reduction at 3 and 6 months postoperatively. The association was not statistically significant at 12 months postoperatively (β=-0.07, [95% CI: -0.24- 0.09], *P* = .54). Various portal functions, including messaging, visits, my record, medical tools, billing, resources, and others, were positively associated with %BMI reduction at 3- and 6-months follow-ups.

**Conclusions:** Greater patient portal engagement, which may represent stronger adherence to postoperative instructions, better self-management of health, and enhanced communication with care teams, was associated with improved postoperative weight loss. Future investigations are needed to identify important portal features that contribute to the long-term success of weight loss management.

## Introduction

Obesity continues to be a significant health concern in the United States, with approximately 42% and 9.2% of adults classified as having obesity (body mass index (BMI) ≥30 kg/m2) and severe obesity (BMI ≥40 kg/m2), respectively.[1] Research has consistently demonstrated that individuals with obesity face an elevated risk for a range of health complications, including but not limited to, cardiovascular disease, diabetes, respiratory problems, and metabolic syndrome.[2– 6] Bariatric surgery remains the most effective intervention for individuals who struggle to achieve sustainable weight loss solely through diet and exercise, whether through its effects of restricting food intake or modifying the digestive system.[7,8]

Comprehensive postoperative care following hospital discharge plays a pivotal role in achieving optimal outcomes and enduring success for bariatric surgery patients.[9] Effective health tracking, patient self-management, and efficient collaboration between patients and their care teams, are of utmost importance in working towards shared goals, including sustained weight loss, improved health conditions, and enhanced overall quality of life. Such postoperative needs have been increasingly supported by both web-based and mobile app-based patient portals,[10] which typically provide patients access to their own electronic health information and serve as platforms for timely communication and regular health information exchange with their care teams. Growing evidence underscores the efficacy of portal usage in delivering continuous support and guidance, monitoring key indicators related to primary interventions, enhancing medication adherence, and fostering self-management in promoting behavioral and lifestyle modifications.[11,12] Despite the reported merits of patient portal usage in general settings of care, its application to bariatric surgery has not been evaluated. In particular, there remains a gap in understanding the link between patient portal engagement and weight loss following bariatric surgery.

In the present study, we investigated the association between patient portal engagement and weight loss outcomes, spanning from 3 months to one-year post-bariatric surgery. We hypothesized that greater engagement with patient portals is associated with enhanced weight loss. Moreover, we investigated how such an association varies over time.

## Methods

This study was approved by the Institutional Review Boards (IRB) at Vanderbilt University Medical Center (VUMC) under IRB#221459. A full waiver of written informed consent from patients was granted by the IRB due to the fact that this study is retrospective with minimal risks to patients. This study follows the Strengthening the Reporting of Observational Studies in Epidemiology (STROBE) reporting guideline.

### Study Settings

We conducted a single-site retrospective longitudinal study at Vanderbilt University Medical Center (VUMC), a large academic medical center in Nashville, TN, USA that provides primary and specialty referral care to patients from across the Southeast. This study included patients who underwent Roux-en-Y gastric bypass (RYGB) or sleeve gastrectomy (SG) operations, the most commonly performed operations,[13] between January 1, 2018, and March 1, 2021, inclusive, as part of their participation in the VUMC Surgical Weight Loss Program. The specific end date reflected the most current data update within the available dataset. The VUMC Surgical Weight Loss Program is accredited by the Metabolic and Bariatric Surgery Accreditation and Quality Improvement Program (MBSAQIP).[14] VUMC deployed its patient portal, My Health at Vanderbilt (MHAV), in 2004, and migrated to Epic’s MyChart platform in late 2017 as part of a wider electronic health record system update across VUMC. Like most patient portals, MHAV allows patients to access their own electronic health information, make appointments, manage medications, and interact with their care providers through a secure messaging system.[15] MHAV currently has over 1 million users and is accessed over 30 million times annually.

### Data

The cohort was obtained from the VUMC bariatric surgery Quality, Efficacy, and Safety (QES) registry, a VUMC-specific database used exclusively for internal research and quality enhancement. We considered initial RYGB or SG operations performed at VUMC and excluded any subsequent surgical revisions due to their inherent complexity. We identified the date of each patient’s initial bariatric surgery (day zero) as the index event. Following the surgery, patients were scheduled for clinic follow-ups at 3, 6, and 12 months postoperative. We collected BMI data from QES registry and EHR. The observations with missing weights at the follow-up visits at the 3, 6, or 12 months post-surgery and the patients who lacked weight records immediately prior to surgery (baseline) were excluded.

Patient demographic and clinical information were extracted from Epic electronic health record (EHR) system, which was deployed at VUMC in 2017. Demographic information includes age at surgery, sex (female or male), and self-reported race and ethnicity (Black, White, or other Races/Ethnicities) from EHR. We grouped American Indian or Alaska Native, Asian, Asian Indian, Chinese, Cuban, Filipino, Guamanian or Chamorro, Hispanic or Latino, Japanese, Korean, Mexican, Mexican American or Chicano, Native Hawaiian, other Asian, other Pacific Islander, Puerto Rican, Samoan, Vietnamese, and none of the above into “other Races/Ethnicities” to avoid unstable estimates due to their small cohort sizes. Past medical conditions of each patient, encompassing a period of 10 years up to the index event, were extracted to calculate the Charlson comorbidity index (CCI).[16,17] To account for socioeconomic status, we used the area deprivation index (ADI),[18] a widely adopted measure indicating the neighborhood socioeconomic disadvantage level. In our analysis, we determined patients’ ADI by their respective 5-digit Zip Codes.

We extracted patient portal engagement history from MHAV event logs, which record every action taken by system users through provided interfaces.[19–24] These included unique patient identifiers, event timestamps, and event types. For each patient, we focused on the timeframe starting from day 5 post-surgery— typically regarded as the commencement of the post-discharge phase—up to 12 months postoperative. We categorized event types to seven portal functions: messaging (support communication between healthcare providers and patients), visits (facilitate appointment management), my record (provide lists of allergies, immunizations, medical history, medications, pharmacy, preventive care, test results, and vitals), medical tools (allow patients to view document and add devices), billing (support account payment and insurance management), resources (provide patient education materials), and others (include additional functions not specified above, such as “Send proxy invite”).[19] The full list of event types associated with these functions is reported in Supplement eTable 1.

### Measure Definition

There is no universally agreed upon measure of patient portal engagement. For this study, we quantified engagement by measuring the number of days in which a patient performed any action within MHAV after logging in, as recorded in the event logs of MHAV. Days that solely consisted of login and logout events were not counted. As such, we use “active days of portal use” interchangeably to refer to this measurement. We trimmed outliers where the number of active days of portal use surpassed 110 (95th quantile of portal use) to mitigate the risk of deriving biased analysis. We tracked the number of active days of portal usage from day 5 to the conclusion of the 3, 6, and 12 postoperative months, respectively.

Postoperative weight loss outcome is defined as the percentage of BMI (%BMI) reduction at month m compared to the baseline BMI:

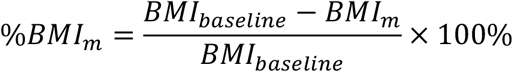

The primary goal of this study is to examine the association between patient portal engagement and %BMI reduction at different follow-up time points.

### Statistical Analysis

The data analysis was performed between February 2023 and November 2023. We used generalized estimating equation [25] with identity link function and exchangeable correlation structure to model the association of patient portal engagement with %BMI reduction. In addition to P-values, effect size estimates were also reported using a robust effect size index (RESI) along with its 95% confidence intervals (CIs).[26,27] The RESI is equal to ½ Cohen’s d under some assumptions,[28] so Cohen’s suggested interpretations for ES are: none to small (RESI=[0, 0.1]); small to medium (RESI=(0.1, 0.25]); and medium to large (RESI=(0.25, 0.4]). The direction of the RESI estimates indicates the association direction. The independent and dependent variable are the number of active days of portal use (winsorized at 110-day) and the %BMI reduction at the 3, 6, or 12 months post-operation, respectively. Covariates included the duration since surgery (in months), the patient’s age at the time of surgery (in years), gender (female or male), race and ethnicity (Black, White, or other Races/Ethnicities), type of bariatric surgery (RYGB or SG), CCI score, and ADI. Conceptual framework was included in Supplement eFigure 2.

We centered our analyses around the primary hypothesis that greater patient portal engagement is associated with more %BMI reduction at a given month, and this association varies over time. Therefore, the primary analysis incorporated the interaction term between portal engagement and duration since surgery. We also conducted a secondary analysis to test the relative influence of specific portal functions. As part of this, we restricted our analysis to the active days of each portal function separately. A sensitivity analysis was performed by adding BMI immediately prior to surgery (BMI at baseline) to the primary model to account for its potential mediation effect. In addition, considering the overlap of our study period with the COVID-19 pandemic, during which MHAV usage spiked as a lot of healthcare moved to virtual delivery,[29] we conducted an additional sensitivity analysis by dividing the patient cohort based on whether their surgeries occurred before March 1, 2020. We then determined if the associations between portal engagement and %BMI reduction were consistent before and during the pandemic.

All statistical analyses were conducted using R software (version 4.3.1). A *P*-value less than 0.05 was considered statistically significant.

## Results

### Descriptive Analysis

The bariatric surgery cohort comprises 1,415 patients, with 3,377 observations within one-year follow-up visits (study cohort flow diagram shown in Supplement eFigure 1). 805 (56.9%) patients underwent RYGB and 610 (43.1%) underwent SG. 1,145 (80.9%) patients are female (Table 1). The mean (SD) age as of the surgery date is 44.5 (11.4) years, with a mean (SD) baseline BMI of 47.0 (7.71) kg/m2. After surgery, 95.1%, 76.7%, and 66.9% of patients had a follow-up at the 3, 6, and 12 months, respectively. The mean (SD) %BMI reduction at the 3, 6, and 12 months is 15.9% (3.73%), 24.1% (5.58%), and 31.1% (8.30%), respectively. Over 98% of bariatric surgery patients utilized MHAV after surgery. Patients have a mean (SD) engagement of 24.2 (13.5), 37.8 (22.9), and 61.0 (41.2) active days within the first 3, 6, and 12 months post-surgery, respectively. Winsorization of portal engagement affected 137 (4.06%) observations from 130 (9.19%) patients.

**Table 1.** Characteristics of study cohort.

| Characteristics | Overall<br>(N=1,415) | RYGB<br>(N=805) | VSG<br>(N=610) |
| --- | --- | --- | --- |
|  | Patient, No. (%) |  |  |
| **BMI records missing** |  |  |  |
| 3 months | 70 (4.9%) | 34 (4.2%) | 36 (5.9%) |
| 6 months | 329 (23.3%) | 172 (21.4%) | 157 (25.7%) |
| 12 months | 469 (33.1%) | 237 (29.4%) | 232 (38.0%) |
| **Gender** |  |  |  |
| Female | 1145 (80.9%) | 656 (81.5%) | 489 (80.2%) |
| Male | 270 (19.1%) | 149 (18.5%) | 121 (19.8%) |
| **Race and ethnicity** |  |  |  |
| Black | 282 (19.9%) | 110 (13.7%) | 172 (28.2%) |
| White | 1088 (76.9%) | 668 (83.0%) | 420 (68.9%) |
| Other Races/Ethnicities | 45 (3.2%) | 27 (3.4%) | 18 (3.0%) |
|  | **Mean (SD)** |  |  |
| **Age at baseline** | 44.5 (11.4) | 44.4 (11.0) | 44.8 (12.0) |
| **Baseline BMI** | 47.0 (7.71) | 47.3 (7.88) | 46.6 (7.48) |
| **%BMI reduction** |  |  |  |
| 3 months | 15.9 (3.73) | 16.5 (3.83) | 15.0 (3.42) |
| 6 months | 24.1 (5.58) | 25.5 (5.21) | 22.0 (5.46) |
| 12 months | 31.1 (8.30) | 33.5 (7.42) | 27.4 (8.22) |
| **Portal active days<sup>a</sup>** |  |  |  |
| 3 months | 24.2 (13.5) | 24.6 (13.9) | 23.7 (12.8) |
| 6 months | 37.8 (22.9) | 38.9 (24.0) | 36.3 (21.4) |
| 12 months | 61.0 (41.2) | 63.2 (43.3) | 58.1 (38.0) |
| **Function active days at 12-months<sup>1</sup>** |  |  |  |
| Billing | 19.5 (17.3) | 20.4 (18.6) | 18.4 (15.4) |
| Medical tools | 38.9 (42.3) | 40.9 (44.7) | 36.2 (38.8) |
| Messaging | 36.9 (25.6) | 37.8 (26.2) | 35.8 (24.7) |
| My record | 59.4 (40.4) | 61.5 (42.4) | 56.7 (37.3) |
| Resources | 9.09 (19.2) | 8.74 (19.2) | 9.56 (19.0) |
| Visit | 55.6 (40.3) | 57.1 (42.2) | 53.6 (37.6) |
| Others | 13.0 (15.1) | 13.9 (16.3) | 11.8 (13.4) |
| **Area Deprivation Index (ADI)** | 56.3 (19.1) | 56.7 (19.3) | 55.7 (18.8) |
| **Charlson Comorbidity Index (CCI)** | 1.23 (1.59) | 1.27 (1.51) | 1.19 (1.68) |
| <sup>a</sup> The portal active days are not winsorized. |  |  |  |

### Multivariate Analysis

Our results revealed significant associations between patient portal engagement and %BMI reduction: an additional 10-day increment in active portal engagement is associated with a 0.57 ([95% CI: 0.42-0.72], *P* < .001), and 0.35 ([95% CI: 0.22-0.49], *P* < .001) increase in the expected %BMI reduction at the 3- and 6-month follow-ups, respectively (Figure 1). This association is not statistically significant at the 12-month follow-ups (β=-0.07, [95% CI: -0.24-0.09], *P* = .54). Moreover, the analysis demonstrated that the effect of portal engagement over %BMI reduction has significant variation over time. For each additional month, the effect over %BMI reduction decreases by 0.07 ([95% CI: -0.09- -0.05], *P* < .001) (Table 2).

**Figure 1.**
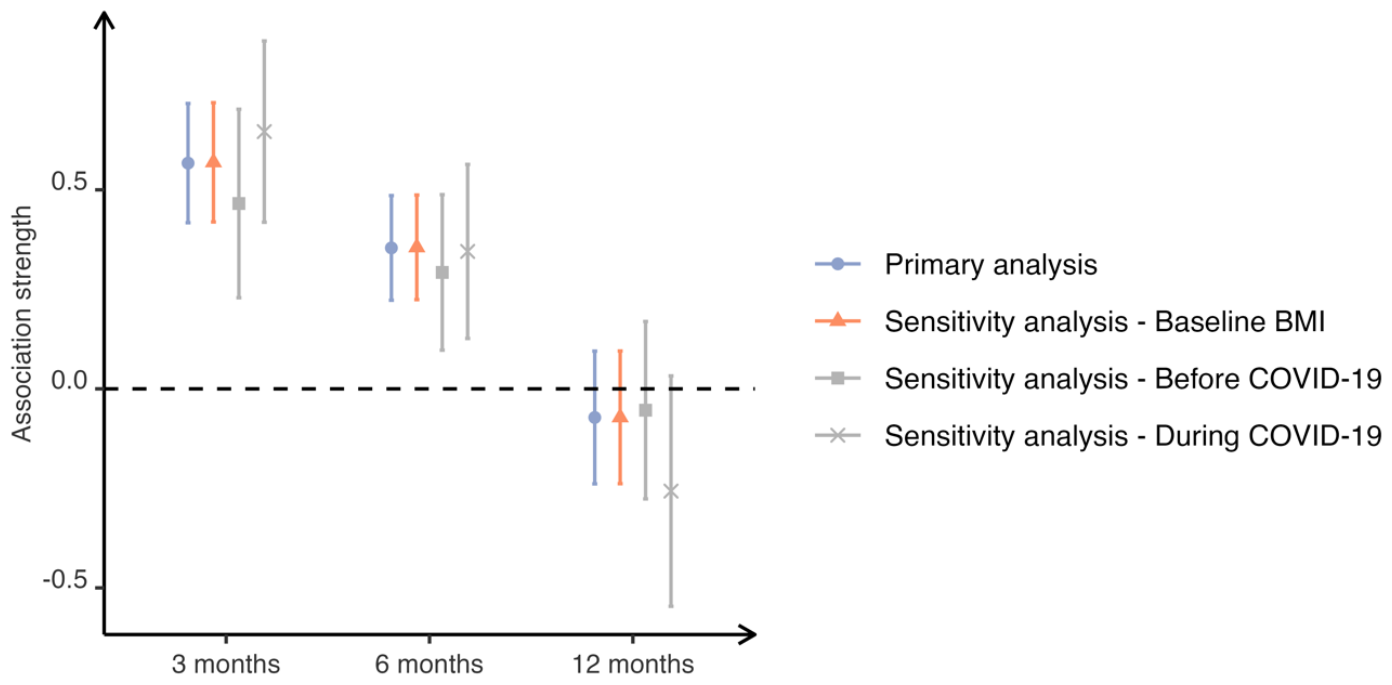
The estimated association strengths (regression coefficients) and the corresponding 95% confidence intervals between portal engagement and the %BMI reduction at the 3, 6 and 12 months, respectively.

**Table 2:** Model results of the relationship between portal engagement and %BMI reduction over time following bariatric surgery. All other independent variables factored in as covariates.

| Independent variables | Regression coefficient (95% CI) | P | RESI (95% CI) |
| --- | --- | --- | --- |
| Portal engagement (per 10-days) | 0.78 [0.62- 0.94] | <.001 | 0.25 [0.19- 0.33] |
| Portal engagement x months | -0.07 [-0.09- -0.05] | <.001 | -0.22 [-0.28- -0.16] |
| Age (per 5-years) | -0.26 [-0.38- -0.14] | <.001 | -0.11 [-0.17- -0.06] |
| Male<sup>a</sup> | 0.21 [-0.41- 0.83] | .51 | 0.02 [-0.04- 0.06] |
| Black<sup>b</sup> | -2.52 [-3.16- -1.87] | <.001 | -0.20 [-0.26- -0.15] |
| Other Races/Ethnicities<sup>b</sup> | -1.54 [-3.19- 0.10] | .07 | -0.05 [-0.11- 0.01] |
| ADI | 0.01 [-0.01- 0.02] | .31 | 0.03 [-0.03- 0.08] |
| CCI | -0.35 [-0.52- -0.18] | <.001 | -0.10 [-0.16- -0.06] |
| SG<sup>c</sup> | -2.90 [-3.41- -2.40] | <.001 | -0.30 [-0.36- -0.24] |
| Months post-surgery | 1.86 [1.76- 1.97] | <.001 | 0.91 [0.84- 1.00] |

<sup>a</sup> The reference group for gender is female. <sup>b</sup> The reference group for race and ethnicity is White. <sup>c</sup> The reference group for operation type is RYGB.

A significantly lower expected %BMI reduction was observed in Black patients (β=- 2.52, [95% CI: -3.16- -1.87], *P* < .001) compared to White patients (Table 2). Such difference is not significant for patients who were self-identified as other Races/Ethnicities (β=-1.54, [95% CI: -3.19- 0.10], *P* = .07). In terms of age at operation, a 5-year increase was found to be significantly associated with a 0.26 decrease in the expected %BMI reduction (β=-0.26, [95% CI: -0.38- -0.14], *P* < .001). When evaluating comorbidities, our findings indicated that patients with a higher CCI score were associated with a significantly lower expected %BMI reduction (β=- 0.35, [95% CI: -0.52- -0.18], *P* < .001). In the context of surgery type, our results indicated the expected %BMI reduction among the patients who underwent SG is 2.90 lower than those who received RYGB (β=-2.90, [95% CI: -3.41- -2.40], *P* < .001). Additionally, patients from areas with different ADI did not show significant differences in %BMI reduction (β=0.01, [95% CI: -0.01- 0.02], *P* = .31).

Regarding specific portal functions, “My Record” and “Visit” functions – enabling patients to access their medications and test results, and manage their appointments, respectively – were the most frequently utilized (Table 1). Our analysis found that every portal function had a significant positive association with %BMI reduction at 3- and 6-month follow-ups. Specifically, “Resources”, “Others”, and “Messaging” functions showed the strongest associations at 3 months (β=1.31, [95% CI: 0.80-1.82], *P* < .001 for “Resources”; β=1.02, [95% CI: 0.63-1.40], *P* < .001 for “Others”; β=0.86, [95% CI: 0.65-1.08], *P* < .001 for “Messaging”) and 6 months (β=0.84, [95% CI: 0.46-1.22], *P* < .001) for “Resources”; β=0.59, [95% CI: 0.28-0.90], *P* < .001 for “Others”; β=0.55, [95% CI: 0.37-0.73], *P* < .001 for “Messaging” (Table 3). However, no functions showed a significant positive association with %BMI reduction at 12-month follow-up.

**Table 3:**
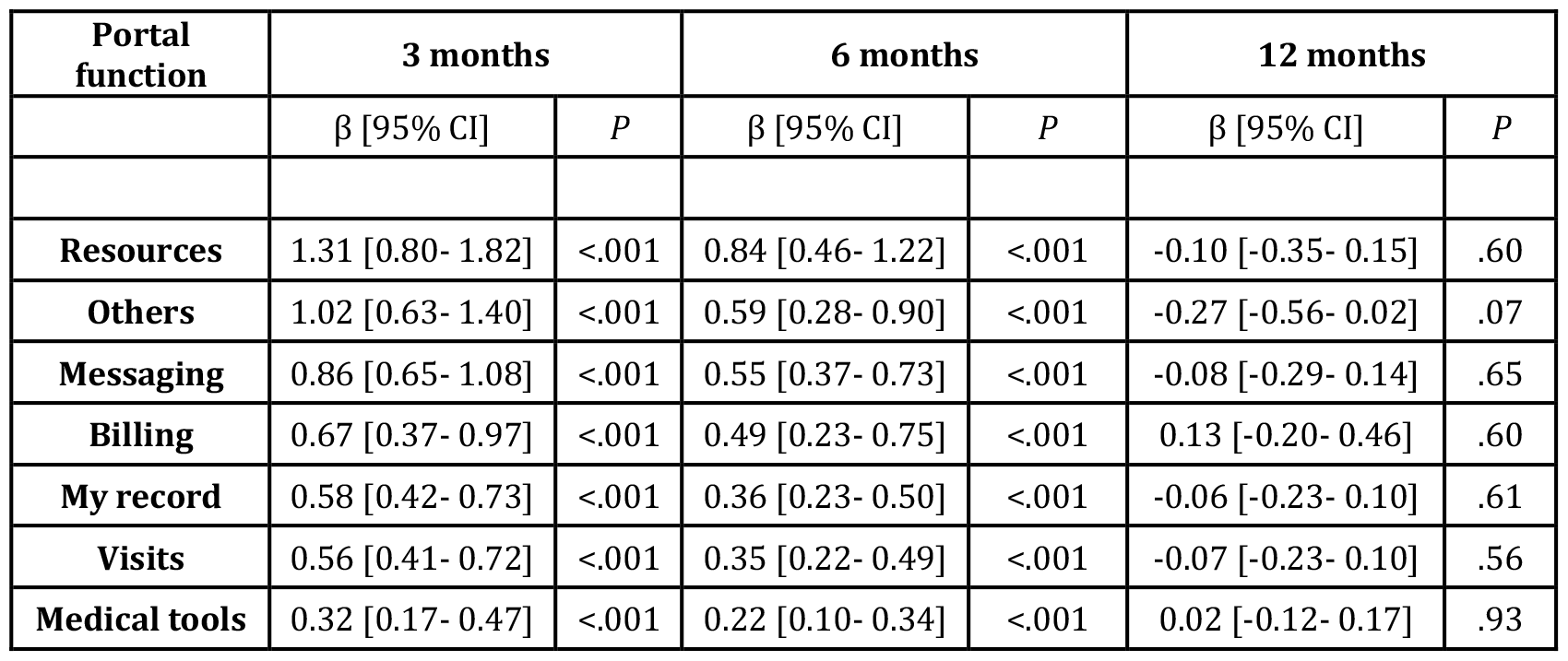
Association strengths between specific portal functions and the expected %BMI reduction at 3, 6 and 12 months.

| Portal function | 3 months |  | 6 months |  | 12 months |  |
| --- | --- | --- | --- | --- | --- | --- |
| | $\beta$ [95% CI] | $P$ | $\beta$ [95% CI] | $P$ | $\beta$ [95% CI] | $P$ |
| **Resources** | 1.31 [0.80- 1.82] | <.001 | 0.84 [0.46- 1.22] | <.001 | -0.10 [-0.35- 0.15] | .60 |
| **Others** | 1.02 [0.63- 1.40] | <.001 | 0.59 [0.28- 0.90] | <.001 | -0.27 [-0.56- 0.02] | .07 |
| **Messaging** | 0.86 [0.65- 1.08] | <.001 | 0.55 [0.37- 0.73] | <.001 | -0.08 [-0.29- 0.14] | .65 |
| **Billing** | 0.67 [0.37- 0.97] | <.001 | 0.49 [0.23- 0.75] | <.001 | 0.13 [-0.20- 0.46] | .60 |
| **My record** | 0.58 [0.42- 0.73] | <.001 | 0.36 [0.23- 0.50] | <.001 | -0.06 [-0.23- 0.10] | .61 |
| **Visits** | 0.56 [0.41- 0.72] | <.001 | 0.35 [0.22- 0.49] | <.001 | -0.07 [-0.23- 0.10] | .56 |
| **Medical tools** | 0.32 [0.17- 0.47] | <.001 | 0.22 [0.10- 0.34] | <.001 | 0.02 [-0.12- 0.17] | .93 |

The sensitivity analysis (Figure 1, Supplement eTable 2) indicated that the positive association between portal engagement and %BMI reduction at both 3 and 6 months persisted after adjusting for baseline BMI (β=0.57, [95% CI: 0.42-0.71], *P* < .001 at 3 months, and β=0.36, [95% CI: 0.22-0.49], *P* < .001 at 6 months). Portal engagement increased significantly during the COVID-19 pandemic, with mean (SD) engagement rising from 51.9 (38.7) active days prior to the pandemic to 77.9 (40.3) days amidst it at 12 months postoperative (Supplement eTable 3). The analysis confirmed that the positive association between portal engagement and %BMI was consistent across the pre-pandemic and pandemic period (Supplement eTables 4 and 5).

## Discussion

Bariatric surgery necessitates sustained postoperative care, and prior studies have suggested that patient portals can enhance health awareness and therapy adherence.[11,30,31] Echoing findings from these studies, our retrospective longitudinal study suggested that increased portal engagement positively associated with better weight loss outcomes post-bariatric surgery with a medium effect size (RESI=0.25, [95% CI: 0.19-0.33]), which underscores the pivotal role of patient portals in potentially enhancing post-surgery care and outcomes.

Our findings emphasized the importance of early patient portal engagement in promoting BMI reduction after bariatric surgery. We demonstrated that the strongest association between portal engagement and BMI reduction presented in the initial 3 months postoperative. This finding aligns with prior studies, which indicated that weight changes during the early postoperative period are predictive of long-term weight loss success.[32–34] Silveria et al. demonstrated that patients achieving less than 5% total body weight reduction at 1 month and 10% at 3 months following RYGB are associated with suboptimal weight loss at 1 year.[33] Manning et al. found that weight loss velocity between 3 to 6 months is associated with maximal weight loss percentage.[34] Our results add to the existing literature by revealing the temporal shifts of patient portal engagement, underscoring the vital role that patient portal engagement plays in fostering favorable long-term weight loss outcomes.

However, loss to follow-ups remains a significant challenge in bariatric surgery studies. In our study, follow-up rates were 95.1%, 76.7%, and 66.9% at 3, 6, and 12 months, respectively. Potentials reasons for this loss includes patients with fewer symptoms opting out of later follow-up appointments at the surgery center, changes in their residence and insurance status, etc [35]. This decline in follow-up rates from 6 months to 12 months may reduce the power to capture the association between portal engagement and BMI reduction and may cause a potential selection bias at 12 months.

Further investigation into the specific functions of the portal revealed that the “Resources” (e.g., “View Education Point”) and “Messaging” functions have the strongest associations with %BMI reduction, suggesting these functions could be particularly beneficial to postoperative management. Notably, by the 6-month follow-up, patients engaging with the “Resources” and “Messaging” functions for every 100 active days saw an additional reduction in BMI by 8.4% and 5.5%, respectively. These findings align with prior study, which indicates that portal may be a source of context-based educational materials for self-management.[12] While “Billing” and “other functions” showed positive associations with %BMI reduction, they frequently co-occurred with other functions. In particular, “Billing” and “other functions” occurred together with “Visit” functions in 92% and 90% active days, respectively, suggesting that their statistical significance may be an artifact of metric design rather than evidence of clinically meaningful postoperative patient engagement through the portal.

Our research provides in-depth, longitudinal analysis of patient portal use in bariatric surgery, highlighting its benefits in the management of postoperative weight loss resources and the communication between clinicians and patients. However, more needs to be done to familiarize both patients and healthcare professionals with portal usage. Encouragement from healthcare providers and user-friendly designs from vendors are essential.[36–38] Nonetheless, the increasing use of portals may lead to clinician burnout,[39,40] underscoring the need for strategies that balance efficient digital interaction management with healthcare team well-being. Achieving this equilibrium is crucial for maximizing patient satisfaction and the effective use of digital health tools.

In addition to patient portal engagement, we observed that younger patients tended to achieve better postoperative outcome (Table 2). This could be attributed to faster metabolic rates in younger individuals.[41] Our findings align with prior studies that reported racial differences in weight loss after bariatric surgery, noting that self-identified Black patients experienced less weight loss compared to their White counterparts; however, the underlying causative reasons remain unclear.[42] We also found that patients with multiple comorbidities exhibited less weight loss, highlighting the needs for tailored postoperative care for patients with more complex health conditions. Our analysis revealed that patient undergoing RYGB lost more weight loss than those who had SG, consistent with existing literature.[8,43] The duration post-surgery was positively associated with BMI reduction. It highlighted the importance of considering the timeline in post-bariatric surgery outcomes analysis. While these observations were significant, they were not the primary focus of this work and warrant further investigation.

## Limitations

Several limitations in this study are worth acknowledging. First, this study was conducted at a single academic medical center. The generalizability of the findings needs to be confirmed in other large surgical weight loss programs. Second, while we adjusted for a range of common factors associated with bariatric surgery outcomes, there could be confounding effects from other factors not within our data access, such as individual-level socioeconomic status and lifestyle factors. Lastly, the direct effects of specific clinic-oriented activities conducted via the portal—such as medication refill requests or appointment scheduling—on improved weight loss outcomes have yet to be determined. In this study, these actions were not investigated in detail due to the smaller sample sizes they represent, which could result in a lack of statistical power and unstable estimates. Moving forward, our research aims to identify and scrutinize these pivotal actions more closely and assess their associations with post-bariatric surgery outcomes.

## Conclusions

In this retrospective study, we examined longitudinal data from VUMC bariatric surgery registry and patient portal utilization. Our findings suggest that higher portal engagement was associated with better post-surgery weight loss. The proposed portal engagement metric and longitudinal analysis framework has the potential to be applied to other diseases that also require long-term care. Future studies can consider incorporating additional post-surgery factors, including dietary intake and comorbidity improvement, to gain a more holistic understanding of long-term outcomes after bariatric surgery.

## Data Availability

The datasets generated and/or analyzed during the current study are not publicly available due to patient private information investigated but are available from the corresponding authors on reasonable request.

## Acknowledgements

XZ, CY, KK, and YC contributed to study conception and design. XZ and YC accessed and verified the underlying data. XZ, KK, CY, YF, and YC performed data analysis and interpretation. KK, XZ, and SV contributed to statistical design. XZ and YC drafted the manuscript. YC obtained funding and supervised this study. All authors contributed to critical revision of the manuscript for important intellectual content.

This research was supported, in part, by the National Institute of Diabetes and Digestive and Kidney Diseases of the National Institutes of Health under Award Number R01DK126721. The content is solely the responsibility of the authors and does not necessarily represent the official views of the National Institutes of Health.

## Conflicts of Interest

GS receives consulting fees from Novo Nordisk, Rhythm Pharmaceuticals, and Eli Lilly. GS is on the Speaker’s Bureau for Novo Nordisk and Rhythm Pharmaceutical and receives industry support for clinical trials from Eli Lilly.

## Notes

### Author Declarations

This study was approved by the Institutional Review Boards (IRB) at Vanderbilt University Medical Center (VUMC) under IRB#221459. A full waiver of written informed consent from patients was granted by the IRB due to the fact that this study is retrospective with minimal risks to patients.

