## Supplement for "Enhanced Patient Portal Engagement Associated with Improved Weight Loss Outcomes in Post-Bariatric Surgery Patients"

### eTable 1: Patient portal functions and event types from Epic's MyChart.[1]

| Portal function | Interaction type |
|---|---|
| **Billing** | Account Payment<br>Hospital Billing Letter Details<br>View Premium Billing Summary<br>Premium Billing Payments<br>Download Premium Billing Invoice<br>Billing Account Summary<br>Opt In to Paperless Premium Billing Invoices<br>Opt Out of Paperless Premium Billing Invoices<br>View Paperless Premium Billing Invoice Status<br>Patient Estimates - Create Estimate<br>Patient Estimates - Estimate Details<br>Patient Estimates - Estimate List<br>View Explanation of Benefits Document<br>Insurance<br>PHR Insurance<br>Update Insurance<br>Insurance ID Card<br>Insurance Card Image Uploaded<br>Insurance Card Image Deleted<br>HB Account Details<br>Hospital Statement Details<br>Account Details<br>Visit Payments<br>Standard Guarantor Payment<br>Account Inquiry<br>Recent Payments<br>Guest Payment<br>Financial Assistance Summary<br>HB/SBO Auto Pay Signup<br>HB/SBO Auto Pay Update<br>Account ReEnable<br>HB/SBO Auto Pay Terminate<br>Benefit Details<br>Quick Pay |
| **Medical tools** | View/Download Lucy CCD<br>Download Visit Summary<br>Download Requested ROI Record<br>Test Result PDF Downloaded<br>Research Studies<br>Generated Share Everywhere Token<br>Redeemed Share Everywhere Token<br>Wallet Card<br>Health Plan Access Control<br>Patient Notes<br>View Clinical Notes |

| | |
|---|---|
| | View Document<br>eSign Document<br>Address Change Request<br>API Clinical Document Access<br>Onboarding Viewed<br>Explore More Card Click<br>Custom Configured Report<br>Change Paperless Status<br>View Scan<br>Patient Photo<br>Device - Add<br>Change Shortcuts - Load Activity<br>COVID-19 Barcodes Generated<br>Request Lucy CCD<br>Create External Link<br>Device - Remove All<br>Implanted Devices<br>Device - Auth<br>Device - List<br>Device - Remove<br>Lab Requisition PDF Generated |
| **Messaging** | Letters<br>Messaging<br>Referral Request<br>Customer Service Request<br>Medical Advice Request<br>Healthplan Request<br>Conversation List<br>Conversation<br>Conversation New Message<br>Conversation Action<br>ROI Record Request<br>EHI Export Request<br>Prescription Message<br>Bedside Messaging Request<br>Message Menu Action |
| **My record** | Allergies<br>PHR Allergies<br>Update Allergies<br>PHR Health Issues<br>Flowsheet Reports List<br>Flowsheet Report Details<br>Patient Entered Flowsheet<br>Health Summary<br>Immunizations<br>PHR Immunizations<br>Update Immunizations |

| | |
|---|---|
| | Medications<br>Medication Renewal Request<br>Update Medications<br>Medication Ingredients<br>Medication reference viewed<br>My Conditions<br>Preventative Care Scheduling<br>Test Result Details<br>Test Results List<br>Medical Histories<br>Problem List<br>Demographics<br>Pharmacies<br>Infectious Disease Status<br>Travel History<br>Result Component Graphing<br>Update Wait List<br>View To Do Changes<br>View Requested ROI Records<br>Health Maintenance - Mark As Complete<br>Community Management<br>Procedures<br>View Visit Summary<br>Transmit Visit Summary<br>Patient Assigned Tasks<br>Growth Chart<br>Update Problems<br>Alerts Retrieved from Remote System<br>Patient Initiated Questionnaires<br>Genetic Profile<br>Health Advisory Reminder Hidden From Health Feed<br>COVID-19 Patient Reconciled Vaccinations<br>COVID-19 Patient Registry Query<br>Immunization Details<br>COVID-19 Registry Immunizations Added<br>COVID-19 Vaccination Record PDF Generated<br>Health Advisory Reminder Unhidden From Health Feed<br>Health Feed<br>COVID-19 Health Card Generated<br>Get PCP<br>Provider List Widget<br>Goals<br>Provider Details<br>Advanced Care Planning<br>Manage Reminders<br>Trends Dashboard<br>Provider Search<br>MyChart Patient Created Tasks |

| | |
|---|---|
| | Health Maintenance<br>Upcoming Orders |
| **Resources** | View Education Title<br>View Education Point<br>Update Education Status<br>Inline Education External<br>Education Question<br>Inline Education Internal<br>Get Patient Prescribed Education Titles<br>Get One Patient Prescribed Education Title<br>Explore More<br>Health Feed Card Clicked<br>Call Center List |
| **Visit** | Appointment Request<br>Appointment Cancel<br>Appointment Confirm<br>Upcoming Appointment Details<br>Appointment Direct Cancel<br>Appointment Snapshot PDF<br>Appointment Offer<br>Add Favorite Appointment<br>Remove Favorite Appointment<br>Use Favorite Appointment<br>Driving Directions<br>eCheck-In<br>Telemedicine Connect<br>Telemedicine Streaming Status Change<br>Telemedicine Disconnect<br>Telemedicine Hardware Test<br>Telemedicine Initialize<br>Encounter Review<br>Visits<br>Request Visit Summary<br>Direct Scheduling Process<br>Inpatient Admissions<br>Scheduling View Event<br>Add To Calendar<br>Encounter Details<br>Scheduling and Other Preferences<br>On My Way<br>Open Scheduling (Internal)<br>Open Scheduling (External)<br>Referral Review<br>Self-Triage Landing Page<br>Self-Triage<br>Geolocation Self Arrival<br>ED Self-Registration |

| | |
|---|---|
| | Past Self-Triage History Page<br>L&D Pre-Admission Created<br>Visit Contact Information<br>User-initiated Self-Arrival Without Location Tracking |
| **Others** | Questionnaire<br>History Questionnaire<br>Other<br>FDI Link Generated<br>Switch Proxy Context<br>Change Password<br>OAuth2 Access Token Refresh<br>Switch Locale<br>Proxy Context Enter<br>Known Device Used<br>Secondary Identity Validation<br>OAuth2 Access Token Generation<br>Unknown Device Used<br>Proxy Context Exit<br>Accept Terms and Conditions<br>Announcement URL Launched<br>Proxy Access (View)<br>Community Authorization<br>Terms and Conditions<br>L&D Pre-Admission Accessed<br>OAuth2 Authorization Code Generation<br>Demographic Authentication<br>Audit Trail<br>Inline External Jump<br>Inline Education Link<br>Full External Jump<br>OAuth2 Token Review<br>View Feature Library Page<br>Confirmed Proxy Invite<br>Secure Email Changed<br>Self-Triage Terms and Conditions<br>Community Link Validation Success<br>Sent Proxy Invite<br>Change Shortcuts - Save User Selected Shortcuts<br>Secure Mobile Phone Changed<br>Password Reset Question Answer<br>Opt In to Two-Factor<br>Form - Proxy Access<br>Declined Terms and Conditions<br>Other - Non Chargeable<br>Revoke Proxy Access<br>View CE Authorization<br>Community Link Validation Failure |

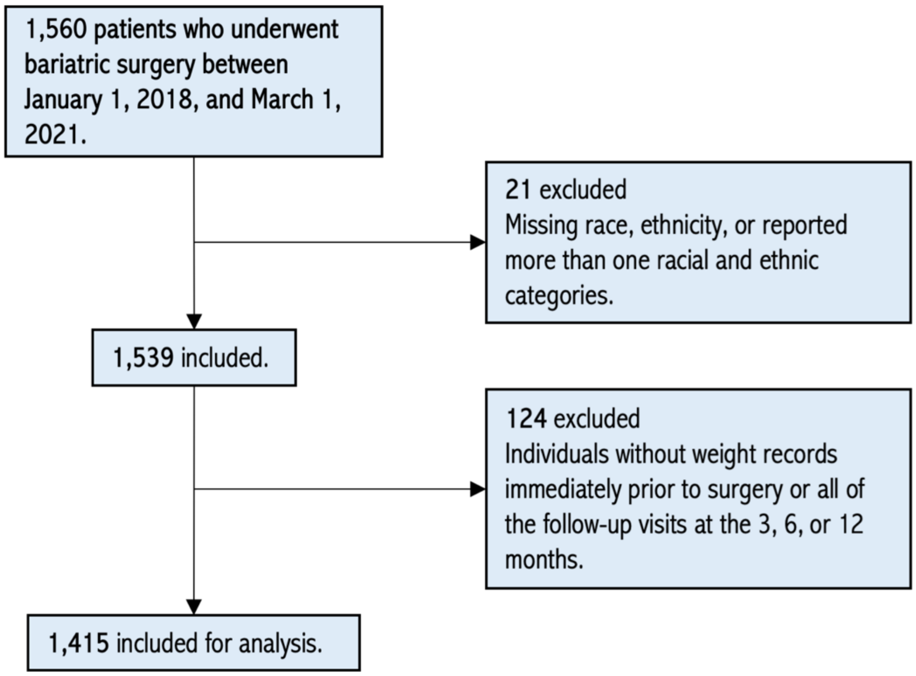


**eFigure 1: Study cohort flow diagrams.** For this study, we removed bariatric surgery patients if they 1) were associated with more than one reported racial or ethnic category or did not have racial or ethnic information recorded or 2) did not have weight records before surgery or all of the follow-up visits at the 3, 6, or 12 months.

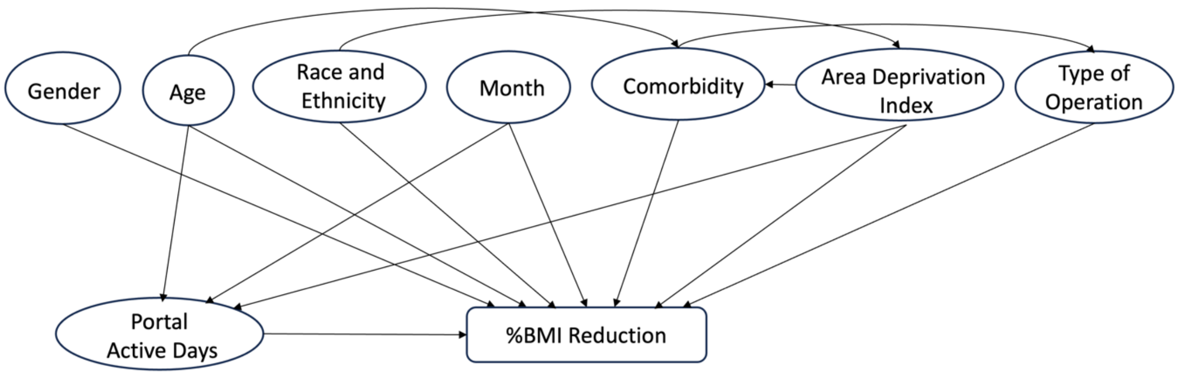


**eFigure 2: Conceptual framework of factors associated with BMI reduction after bariatric surgery.** The primary exposure variable is the number of days patients used patient portal after bariatric surgery, and the dependent variable is the percentage decrease in BMI (%BMI) at 3 months, 6 months, and one year following the bariatric surgery. We incorporated gender, age, race and ethnicity, month after surgery, comorbidity, Area Deprivation index, and type of operation as covariates. We reviewed previous literature[2-6] to identify the associations among non-dependent variables.

**eTable 2:** Model results of the sensitivity analysis – baseline BMI. This model accounts for baseline BMI as an independent variable, complemented by the full set of covariates utilized in the primary analysis model.

| Independent variables | Regression coefficient [95% CI] | *P* | RESI [95% CI] |
|---|---|---|---|
| **Baseline BMI** | -0.06 [-0.09, -0.03] | <.001 | -0.09 [-0.15- -0.04] |
| **Portal engagement (per 10-days)** | 0.78 [0.62, 0.94] | <.001 | 0.25 [0.19- 0.33] |
| **Portal engagement x months** | -0.07 [-0.09, -0.05] | <.001 | -0.22 [-0.28- -0.16] |
| **Age (per 5-years)** | -0.31 [-0.43, -0.19] | <.001 | -0.14 [-0.20- -0.08] |
| **Male[1]** | 0.30 [-0.32, 0.93] | .34 | 0.03 [-0.03- 0.07] |
| **Black[2]** | -2.45 [-3.10, -1.80] | <.001 | -0.20 [-0.25- -0.14] |
| **Other Races/Ethnicities[2]** | -1.55 [-3.17, 0.08] | .06 | -0.05 [-0.11- 0.01] |
| **ADI** | 0.01 [-0.01, 0.02] | .28 | 0.03 [-0.03- 0.08] |
| **CCI** | -0.33 [-0.50, -0.16] | <.001 | -0.10 [-0.15- -0.05] |
| **SG[3]** | -2.94 [-3.45, -2.44] | <.001 | -0.30 [-0.36- -0.25] |
| **Months post-surgery** | 1.86 [1.76, 1.97] | <.001 | 0.91 [0.84- 0.99] |

[1] The reference group for gender is female.
[2] The reference group for race and ethnicity is White.
[3] The reference group for operation type is RYGB.

**eTable 3:** Characteristics of study cohorts based on whether patients' surgery occurred before March 1, 2020, or after.

| Characteristics | Prior to COVID-19 (N=918) | During COVID-19 (N=497) | *P*[1] |
|---|---|---|---|
| | Patient, No. (%) | | |
| **Type of operation** | | | .05 |
| RYGB | 540 (58.8%) | 265 (53.3%) | |
| SG | 378 (41.2%) | 232 (46.7%) | |
| **Gender** | | | .54 |
| Female | 738 (80.4%) | 407 (81.9%) | |
| Male | 180 (19.6%) | 90 (18.1%) | |
| **Race and ethnicity** | | | . 45 |
| Black | 176 (19.2%) | 106 (21.3%) | |
| White | 710 (77.3%) | 378 (76.1%) | |
| Hispanic and Other | 32 (3.5%) | 13 (2.6%) | |
| | Mean (SD) | | |
| **Age at baseline** | 44.9 (11.4) | 43.9 (11.3) | .11 |
| **Baseline BMI** | 47.0 (7.61) | 47.0 (7.91) | .97 |
| **%BMI reduction** | | | |
| 3 months | 15.6 (3.92) | 16.4 (3.29) | <.001 |
| 6 months | 23.8 (5.74) | 24.5 (5.29) | .04 |
| 12 months | 30.8 (8.34) | 31.5 (8.24) | .19 |
| **Portal active days** | | | |
| 3 months | 20.2 (12.6) | 31.7 (11.6) | <.001 |
| 6 months | 31.7 (21.5) | 49.0 (21.3) | <.001 |
| 12 months | 51.9 (38.7) | 77.9 (40.3) | <.001 |
| **Charlson Comorbidity Index** | 1.24 (1.54) | 1.21 (1.68) | .74 |
| **Area Deprivation Index** | 56.1 (19.2) | 56.7 (18.9) | .58 |

[1]Comparison between the before COVID-19 and the during COVID-19 cohort was performed with 2-sample *t* test for numeric variables and Chi-Squared test of independence for categorical variables.

**eTable 4:** Model results of the sensitivity analysis prior to COVID-19. This table presents the model results derived from patient cohort who underwent surgery on or before March 1, 2020.

| Independent variables | Regression coefficient [95% CI] | *P* | RESI [95% CI] |
|---|---|---|---|
| **Portal engagement (per 10-days)** | 0.64 [0.38- 0.90] | <.001 | 0.16 [0.14- 0.31] |
| **Portal engagement x months** | -0.06 [-0.08- -0.03] | <.001 | -0.15 [-0.27- -0.09] |
| **Age (per 5-years)** | -0.23 [-0.38- -0.08] | .003 | -0.10 [-0.22- -0.06] |
| **Male[1]** | -0.09 [-0.85- 0.68] | .82 | -0.01 [-0.04- 0.12] |
| **Black[2]** | -2.16 [-3.01- -1.31] | <.001 | -0.17 [-0.28- -0.10] |
| **Other Races/Ethnicities[2]** | -1.52 [-3.44- 0.40] | .12 | -0.05 [-0.10- 0.07] |
| **ADI** | 0.01 [-0.01- 0.02] | .45 | 0.03 [-0.08- 0.09] |
| **CCI** | -0.30 [-0.52- -0.08] | .008 | -0.09 [-0.21- -0.05] |
| **SG[3]** | -3.28 [-3.92- -2.64] | <.001 | -0.33 [-0.44- -0.27] |
| **Months post-surgery** | 1.80 [1.67- 1.93] | <.001 | 0.92 [0.84- 1.07] |

[1] The reference group for gender is female.
[2] The reference group for race and ethnicity is White.
[3] The reference group for operation type is RYGB.

**eTable 5:** Model results of the sensitivity analysis during COVID-19. This table presents the model results derived from patient cohort who underwent surgery after March 1, 2020.

| Independent variables | Regression coefficient [95% CI] | *P* | RESI [95% CI] |
|---|---|---|---|
| **Portal engagement (per 10-days)** | 0.95 [0.71- 1.18] | <.001 | 0.36 [0.25- 0.69] |
| **Portal engagement x months** | -0.10 [-0.13- -0.08] | <.001 | -0.35 [-0.59- -0.21] |
| **Age (per 5-years)** | -0.31 [-0.50- -0.12] | .001 | -0.14 [-0.33- -0.03] |
| **Male[1]** | 0.78 [-0.25- 1.81] | .14 | 0.07 [-0.01- 0.27] |
| **Black[2]** | -3.17 [-4.15- -2.19] | <.001 | -0.28 [-0.32- -0.04] |
| **Other Races/Ethnicities[2]** | -1.73 [-4.93- 1.47] | .29 | -0.05 [-0.16- 0.34] |
| **ADI** | 0.01 [-0.02- 0.03] | .61 | 0.02 [-0.05- 0.23] |
| **CCI** | -0.39 [-0.67- -0.12] | .005 | -0.13 [-0.37- -0.03] |
| **SG[3]** | -2.33 [-3.15- -1.52] | <.001 | -0.25 [-0.34- -0.05] |
| **Months post-surgery** | 2.10 [1.89- 2.30] | <.001 | 0.91 [0.74- 1.12] |

[1] The reference group for gender is female.
[2] The reference group for race and ethnicity is White.
[3] The reference group for operation type is RYGB.
